# Combining short and long read sequencing technologies to identify SARS-CoV-2 variants in wastewater

**DOI:** 10.1101/2024.08.07.24311639

**Authors:** Gabrielle Jayme, Ju-Ling Liu, Jose Hector Galvez, Sarah Julia Reiling, Sukriye Celikkol Aydin, Arnaud N’Guessan, Sally Lee, Shu-Huang Chen, Alexandra Tsitouras, Fernando Sanchez-Quete, Thomas Maere, Eyerusalem Goitom, Mounia Hachad, Elisabeth Mercier, Stephanie Katharine Loeb, Peter Vanrolleghem, Sarah Dorner, Robert Delatolla, B. Jesse Shapiro, Dominic Frigon, Jiannis Ragoussis, Terrance P. Snutch

## Abstract

During the COVID-19 pandemic, the monitoring of SARS-COV-2 RNA in wastewater was used to track the evolution and emergence of variant lineages and gauge infection levels in the community, informing appropriate public health responses without relying solely on clinical testing. As more sublineages were discovered, it increased the difficulty in identifying distinct variants in a mixed population sample, particularly those without a known lineage. Here, we compare two next-generation sequencing technologies, Illumina and Nanopore, in order to determine their efficacy at detecting variants of differing abundance, using 248 wastewater samples from various Quebec and Ontario cities. Our study used two analytical approaches to identify main variants in the samples: the presence of signature and marker mutations, and the co-occurrence of signature mutations within the same amplicon. We observed that each sequencing method detected certain variants at different frequencies as each method preferentially detects mutations of distinct variants. Illumina sequencing detected more mutations with a predominant lineage that is in low abundance across the population or unknown for that time period, while Nanopore sequencing had a higher detection rate of mutations that are predominantly found in the high abundance B.1.1.7 (Alpha) lineage as well as a higher sequencing rate of co-occurring mutations in the same amplicon. We present a workflow that integrates short read and long read sequencing to improve the detection of SARS-CoV-2 variant lineages in mixed population samples, such as wastewater.

## 1. Introduction

The SARS-CoV-2 genome is constantly evolving, with mutations happening at a rate of about once every 2 weeks [1]. While not all mutations change the characteristics of the virus, some mutations have proven to be of greater concern. Variants of interest (VOI) are labelled as such when an observed lineage is shown to have mutations potentially causing increased transmissibility or virulence, among other attributes [2]. Health organisations may reclassify these variants as variants of concern (VOC) if there is a demonstrable impact on epidemiological data. These viruses are labelled by WHO and assigned a lineage based on PANGO nomenclature [3].

Wastewater surveillance has emerged as a crucial tool in tracking mutations in the SARS-CoV-2 genome. Samples of untreated wastewater can be collected to provide useful information about the spread of COVID-19 in the community, without relying on clinical testing [4,5]. As clinical sampling mainly relies on symptomatic testing, wastewater sampling can provide unbiased and consistent data which can be used to inform appropriate public health responses. It is used to detect variants earlier and provide more context on the transmissibility and COVID-19 levels in communities, particularly where access to clinical testing is not readily available. As wastewater samples consist of a mixture of fragmented RNA from many sources, it can be difficult to accurately identify mutations and variants, particularly those without a known lineage [6].

Next-generation sequencing has proven to be an important tool in pandemic surveillance, particularly in the early detection and spread of variants [7-9]. With a high rate of occurrence of mutations and increased transmissibility, the need to provide high throughput data generation in a relatively short time frame has led to the development of a number of tools and protocols using next-generation sequencing, such as SARS-CoV-2 specific primers and tools to determine lineage in samples. These sequencing methods have been useful in analysing clinical and environmental samples, assisting in tracking viral load, transmission, contact tracing, and virus evolution [8]. Illumina and Nanopore sequencing are two next-generation sequencing technologies that have become major tools in genomic research. Illumina sequencing is a second-generation sequencing technology that uses sequencing by synthesis (SBS), where a reversible fluorescent terminator is used to detect the nucleotide sequence [10,11]. Nanopore sequencing is a third-generation sequencing technology that uses the current changes in a charged protein nanopore from the molecule passing through to determine the specific sequence [10,12]. Multiple studies have been done on the comparison of Nanopore and Illumina sequencing, highlighting their various advantages in different applications [13-15]. Illumina sequencing is widely regarded as being highly accurate, consistently sequencing around 99.5-99.9% accuracy, and the higher depth of reads enables it to be a useful tool in circumstances with poor sequencing coverage, such as wastewater surveillance [16]. Nanopore sequencing has the ability to produce ultra-long reads, only limited by the sample preparation and quality, and is useful in genomic assembly and spanning entire regions of repetitive bases and structural variation [17]. Furthermore, real-time analysis of sequences and portability of sequencing devices has benefits in the field. Studies have been completed comparing Illumina and Nanopore sequencing on clinical and wastewater SARS-CoV-2 samples, which focuses on benchmarking parameters such as genome coverage and depth and variant calling on samples. However, they did not explore the combination of the two sequencing technologies as a method to improve detection of variants [18-20].

In this work, we look to highlight the advantages of both Illumina and Nanopore sequencing in tracking SARS-CoV-2 variants from wastewater samples. Mutational analysis on samples sequenced with both methods allows for a comparison of major variants identified among each dataset.

## 2. Materials and Methods

### 2.1. Sample Collection and Sequencing

Wastewater samples were collected from Ottawa (Ontario, Canada), and Montreal and Quebec City (Quebec, Canada) between March 2020 and March 2021. For Ontario wastewater samples, 24-hour, 500 mL composite primary clarified sludge (PCS) samples were harvested from the City of Ottawa’s Robert O. Pickard Environmental Centre (Ontario, Canada). Samples were transported to the laboratory on ice and immediately processed as described by D’Aoust et al [21]. PCS samples were concentrated by reacting 32 mL of homogenised PCS with a NaCl/polyethylene glycol (PEG) solution at a working concentration of 0.3M NaCl and 80 mg/L PEG, while being agitated for a period of 12-17h at 4°C [22,23]. Afterwards, samples were centrifuged at 10,000 x g for 45 minutes at 4°C, then at 10,000 x g for 10 minutes at 4°C, with the supernatant being discarded after each run. Viral RNA was extracted from the resulting pellet using the RNeasy PowerMicrobiome Kit (Qiagen, Germantown, MD) as per the manufacturer’s instructions with the following exceptions – exactly 250 mg of the resulting pellet was inputted into the extraction kit, and the chloroform-phenom solution was substituted with Trizol LS reagent (ThermoFisher, Ottawa, Canada). Extracted nucleic acids were eluted in 100 μL RNAse and DNAse free water, and frozen at -80°C until further processing.

Montreal and Quebec City wastewater samples were collected by grab sampling, composite sampling, and passive sampling. The composite samples were collected with autosamplers, which collected wastewater every 10 minutes, over a 24-, 48-, or 72-hour time period (depending on the week, day, and sampling sites). The passive samples were collected through 2 absorbent materials, gauze and negatively charged membranes, Mixed Cellulose Ester (MCE) filters, which were housed in torpedoes, also deployed over a 24-, 48-, or 72-hour time period [24]. Grab and composite wastewater samples were additionally concentrated by filtration. Using 50 mL of wastewater, the pH was adjusted to between 3.5 and 4.5, and MgCl2 was added to a final concentration of 25 mM. The samples were then filtered through a 0.45 μm MCE filter. The MCE filters and gauze were stored at -80°C and all samples were processed within 72 hours. RNA was extracted using the Qiagen Allprep Powerviral DNA/RNA kit. The protocol was followed according to the manufacturer, with the exception of the lysis step, where a final concentration of 10% beta-mercaptoethanol was used in the lysis buffer and incubation time was raised to 30 min at 55°C.

RNA extracts of each sample were reverse transcribed with the NEB LunaScript® RT SuperMix Kit and followed by targeted SARS-CoV-2 amplification using the ARTIC V3 primer scheme with NEB Q5 Hot Start High-Fidelity 2X Master Mix or amplicons prepared by Qiagen QIAseq SARS-CoV-2 Primer Panel [25]. Then amplicon PCR products were purified and normalised based on their concentrations. Native barcoded library preparation was performed for Nanopore amplicon sequencing on a PromethION instrument, or a Nextera DNA Flex library preparation was performed for Illumina paired-end amplicon sequencing (PE150) on a NovaSeq 6000 instrument at the McGill Genome Centre. The detailed protocol can be accessed at: https://dx.doi.org/10.17504/protocols.io.by6xpzfn and https://dx.doi.org/10.17504/protocols.io.bmijk4cn.

### 2.2. Mutation Calling and Filtering

For Illumina sequencing data, raw reads were first trimmed using cutadapt (v2.10), then aligned to the reference using bwa-mem (v0.7.17) [26,27]. Aligned reads were filtered using sambamba (v0.7.0) to remove paired reads with an insert size outside the 60-300 bp range, unmapped reads, and all secondary alignments [28]. Then, any remaining primer sequences were trimmed with iVar (v.1.3.4) [29]. Afterwards, a pileup was produced using Samtools (v1.9) which was then used as input for FreeBayes (v1.2.2) to create a consensus sequence and perform variant calling [30,31].

For Oxford Nanopore Technologies (a.k.a. Nanopore) sequencing data, raw signals were basecalled using guppy (v3.4.4) with the High-Accuracy Model (dna_r9.4.1_450bps_hac). Mutation calling for basecalled Nanopore samples was performed through the ARTIC nCoV-2019 pipeline and medaka_haploid_variant workflow (v1.6.0) aligning to the reference genome MN908947.3 [32]. Initial comparison of mutations between the two datasets revealed that 84.1% of mutations across all samples only occurred once in the Nanopore dataset and were not present in the Illumina dataset. To reduce the amount of background error present in the Nanopore sample set, mutations were filtered based on frequency of mutations. Three control samples (1 negative control and 2 positive controls from AccuGenomics [33]) were also sequenced with each method to take into account background noise removal for all samples, due to errors occurring from sample preparation to sequencing. These filters allowed for a more accurate comparison of variants downstream. Coverage analysis to compare read depth and quality was performed with minimap2 and samtools (v1.10) coverage [34,30].

### 2.3. Detection of Variant Lineages in Wastewater Samples

The presence of signature and marker mutations was used to determine the presence of SARS-CoV-2 lineages in the wastewater samples. Signature mutations are those mutations used to define a lineage, taken from PANGO constellations, while marker mutations denote a signature mutation that is only highly prevalent in a certain lineage when compared to other variant lineages [35,36]. To confirm the presence of lineages in the wastewater samples, Freyja was used to determine relative lineage abundances in the dataset [37]. The frequency of variants between Illumina and Nanopore datasets was compared using Barnard’s exact test due to the smaller sample sizes.

To determine the predominant lineage of single mutations, each mutation was searched for using CoV-Spectrum [38]. Using this database, the overall proportion of sequenced samples containing a mutation and the variant lineages of these samples can be established over a set time period. This method was used on mutations that were preferentially detected by a single sequencing method, either Illumina or Nanopore.

Similar to the analysis employed in COJAC, the detection of the co-occurrence of signature mutations within the same amplicon was used to compare the detection of the B.1.1.7 lineage in both datasets [39]. 5 amplicons were found to have 2-4 Alpha signature mutations co-located in a number of samples and were compared to the overall frequency of these mutations. The chi-squared test was used to compare the frequency of co-occurrences between the Illumina and Nanopore datasets.

Python (v3.9.12) was used for all statistical analysis and data visualisation [40]. Scripts relating to mutation and variant analyses can be found in Supplementary Materials.

## 3. Results

### 3.1. Wastewater Sampling and Sequencing with Short and Long Read Methods

We collected 248 wastewater samples from various cities in the provinces of Quebec and Ontario (Canada) and sequenced all samples with both Illumina and Nanopore sequencing. To better understand the differences in variant analyses by both sequencing technologies, we compared read statistics from the samples sequenced of either technology-derived datasets (Table 1). As expected, samples from the Illumina dataset were sequenced at a higher read depth than those from the Nanopore dataset, and overall, the depth of coverage was 562% higher (Figure 1). Average base quality in the Illumina dataset was also higher, with a Q30 score corresponding to a 99.9% accuracy, compared to a Q20 average score for the Nanopore dataset, equating to a 99% accuracy [41]. However, Nanopore sequencing recovered longer reads, allowing them to span the entire amplicon length.

**Table 1.** Comparison of read statistics between Illumina and Nanopore datasets. Statistics are presented with standard deviation and include: the mean number of reads sequenced per sample, the mean read length across all samples, the mean depth of coverage across the SARS-CoV-2 genome for a single sample, and the mean base quality of reads across all samples.

|  | Illumina | Nanopore |
| --- | --- | --- |
| Mean # Reads/Sample | 581,699 ± 66,117 | 21,103 ± 3,653 |
| Mean Length (bp) | 102.1 ± 2.9 | 491.4 ± 14.6 |
| Mean Depth (reads) | 2,345.9 ± 269.7 | 240.9 ± 42.4 |
| Base Quality (Q score) | 30.0 ± 0.8 | 19.9 ± 0.4 |

**Figure 1.**
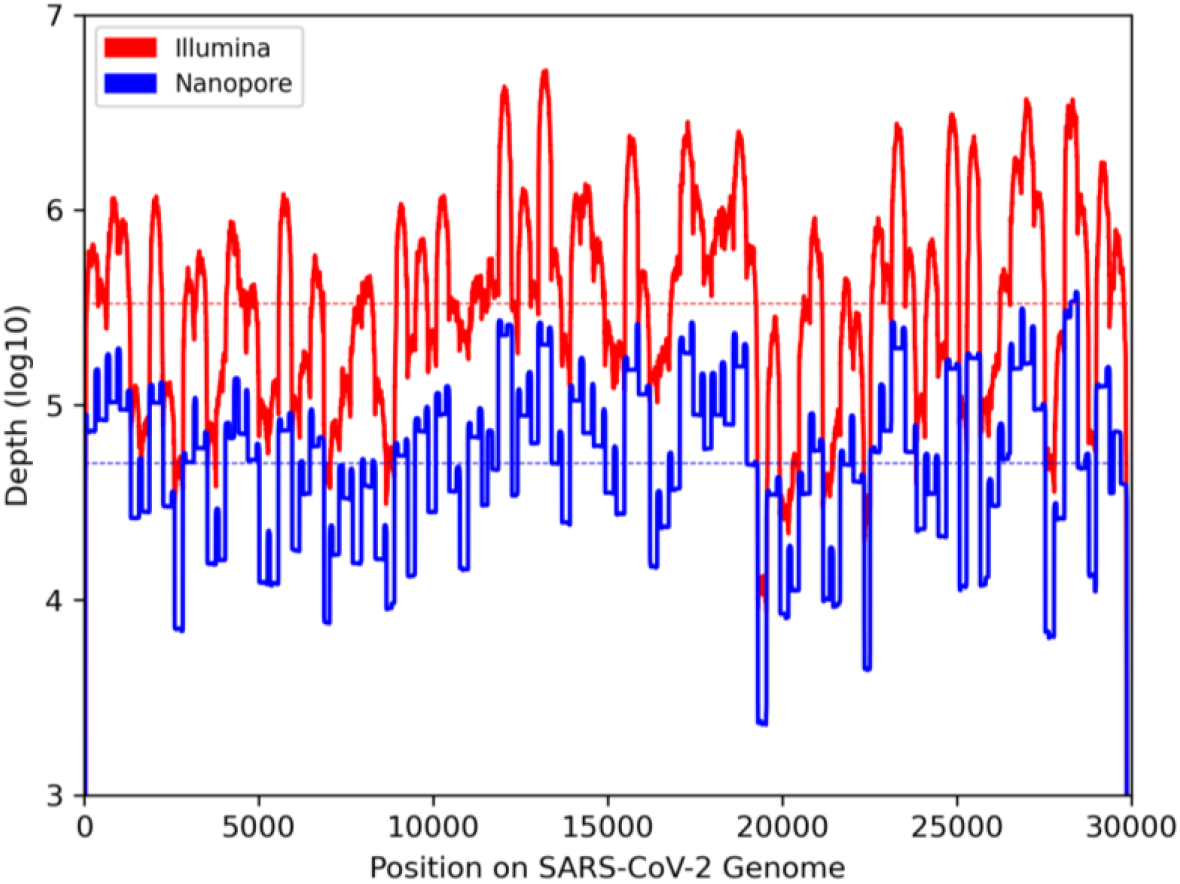
Comparison of depth of coverage across the SARS-CoV-2 genome between Illumina (red) and Nanopore (blue) datasets. Read depth of all wastewater samples within a dataset were combined to show overall coverage on a log10 scale. Horizontal lines indicate average depth across all positions.

### 3.2. Comparison of Mutation Frequency across All Samples

To study the SARS-CoV-2 variants present in the wastewater samples, we performed mutation calling on both datasets (Table S1). Initial analysis yielded 23,688 mutations across all samples, with 94.7% of these mutations exclusive to samples sequenced by Nanopore sequencing (Table 2). Further analysis showed that of the 23,105 total mutations in the Nanopore sample set, 86.3% of them appeared in only one sample, with many more appearing in a low number of samples as well. Given the greater base quality and higher depth of reads in the Illumina dataset, we attributed the high number of Nanopore mutations to background noise and looked to filter these mutations to provide a more accurate comparison of variants in downstream analysis. Due to the lower coverage in individual samples from the Nanopore dataset, filtering mutations on a sample-by-sample basis led to an overcorrection and signature mutations of VOCs were filtered out. Using the procedures described in the Materials and Methods section, filtering was completed on the overall Nanopore sample set to provide a more comprehensive set of mutations to perform variant analysis (Table S2). This gave us a more definitive comparison of the frequency of mutations between Illumina and Nanopore datasets (Figure 2).

**Table 2.** Number of mutations across all SARS-CoV-2 wastewater samples. Counts were recorded after variant calling all samples, as well as after quality filtering the datasets. The number of mutations that are exclusively found within a dataset (Illumina or Nanopore) are also noted, with a significant reduction in exclusively Nanopore mutations that can be attributed to background noise.

| Initial Analysis |  |  |
| --- | --- | --- |
| Number of Mutations |  | Exclusive |
| Total | 23,688 | N/A |
| Illumina | 1,259 | 583 |
| Nanopore | 23,105 | 22,429 |
| After Filtering |  |  |
| Number of Mutations |  | Exclusive |
| Total | 1,259 | N/A |
| Illumina | 1,249 | 534 |
| Nanopore | 725 | 10 |

**Figure 2.**
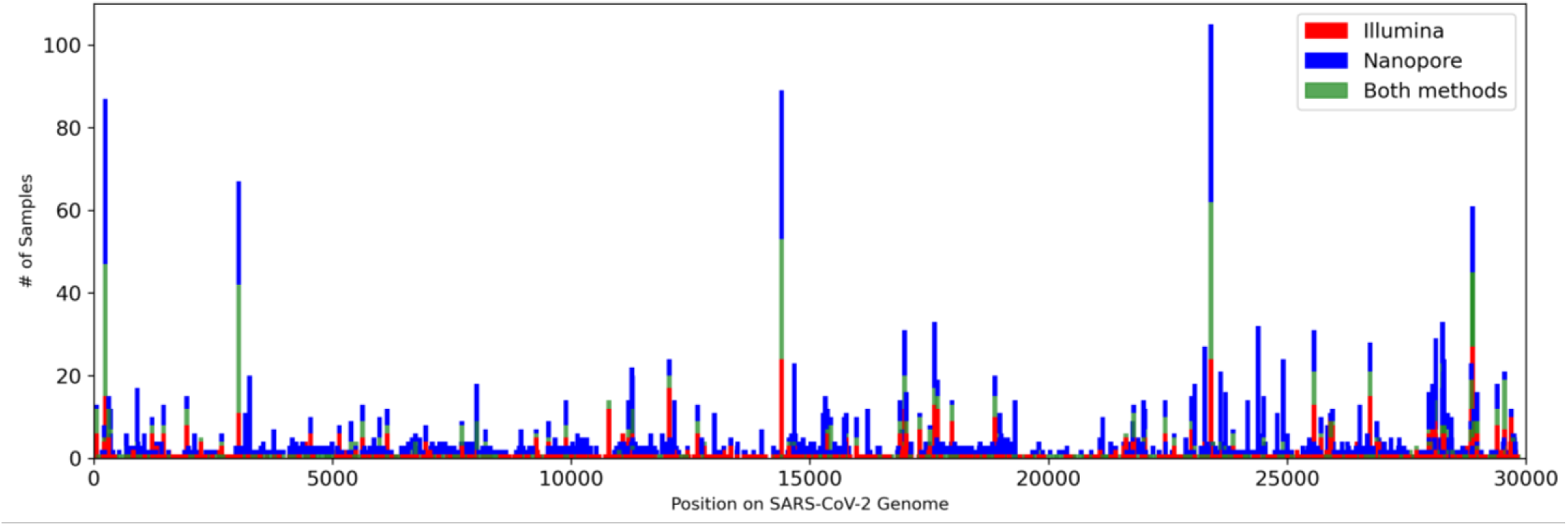
Frequency of mutations across all SARS-CoV-2 wastewater samples. Each bar represents a mutation along the SARS-CoV-2 genome, with red and blue bars indicating the number of samples that exclusively contain that mutation within the Illumina and Nanopore datasets, respectively, and green bars indicating the number of samples in which the mutations occur in both datasets.

### 3.3. Use of Signature and Marker Mutations to Detect Lineages

Using the mutations obtained above, we were able to determine the presence of various lineages in the wastewater samples. First, we sought to identify the variants of concern (VOCs) and variants of interest (VOIs) that were present at the time of sampling. These variants were assigned a lineage using PANGO nomenclature [3]. Using the approach presented by N’Guessan et al., we determined the presence of a major variant in a sample by the occurrence of at least 3 signature mutations and 1 marker mutation [36].

As expected for the time period, the Alpha variant (B.1.1.7) was the most abundant among both Illumina and Nanopore datasets, with an increased identification of variants by combining both datasets together (Figure 3). Of the 9 VOCs/VOIs detected in the overall population, Nanopore identified 5 of them (Alpha, Gamma, Delta, Zeta, Eta), and all in equal or higher frequency compared to the Illumina dataset. Illumina detected all 9 variants across the population, including the 4 (Beta, Theta, Lambda, Mu) not identified by Nanopore. Two variants had a significant difference in frequency between Illumina and Nanopore datasets: Nanopore sequencing detected the Alpha variant in significantly more samples (p = 0.0027), whilst Illumina detected the Beta variant in significantly more samples (p = 0.022).

**Figure 3.**
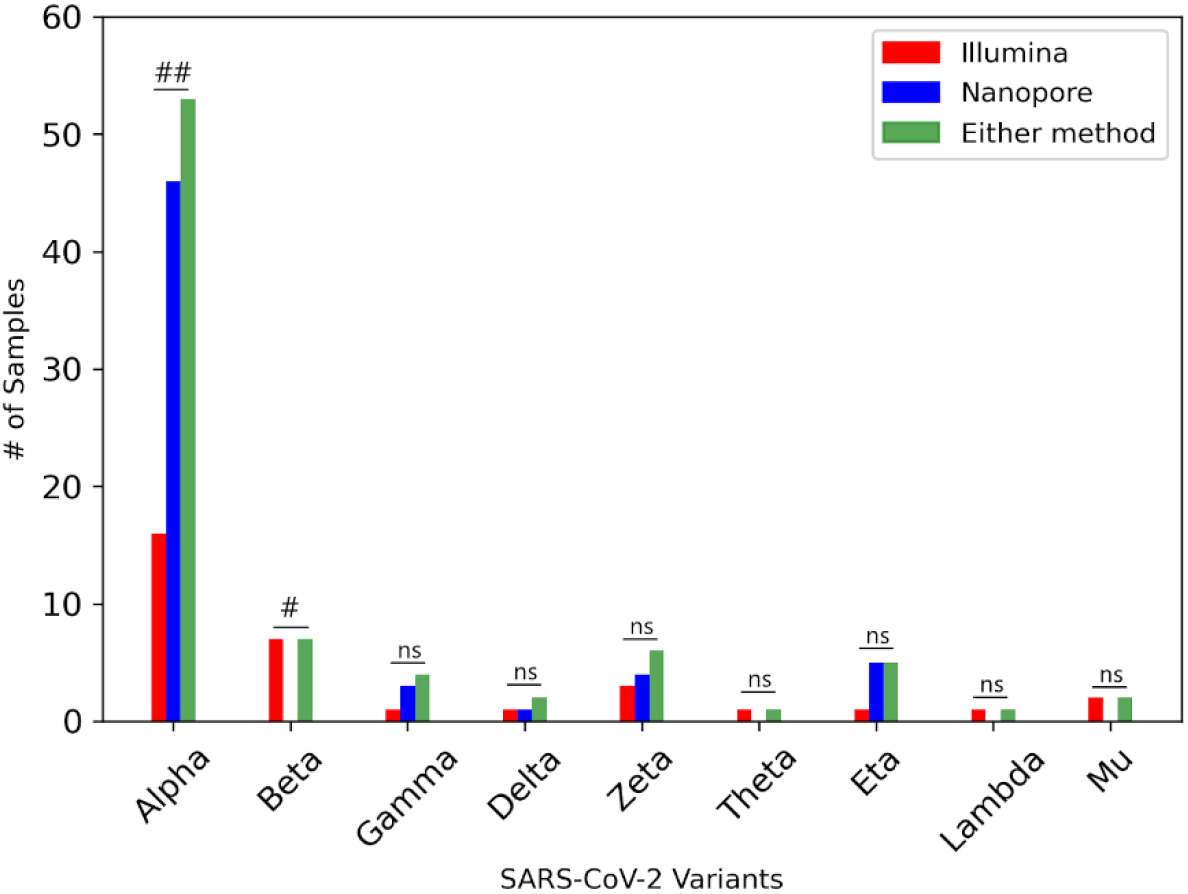
Relative abundance of VOCs/VOIs in SARS-CoV-2 wastewater samples. Using a combination of Freyja and the detection of signature and marker mutations [8], the variants found within each sample were identified. Green bars indicate the number of samples containing each variant, as detected with either sequencing method, compared to variants detected only in the Illumina (red) or Nanopore (blue) datasets. Barnard’s exact test: ##: p < 0.01, #: p < 0.05, ns: p > 0.05.

### 3.4. Predominant Lineages of Key Mutations from Each Dataset

We compared the lineages that were preferentially detected by each sequencing method by looking at the 5 most frequent mutations occurring within a dataset, that were found in low occurrence with the other sequencing method (Table 3). Using CoV-Spectrum, we searched for all lineages in which these mutations are present and identified the most predominant lineage among all consensus sequences at the time of sampling and over time [38].

**Table 3.** Predominant lineages of mutations preferentially detected by a sequencing method. Predominant lineage indicates that of all samples containing the mutation, the indicated lineage is present in the highest percentage of samples.

| Mutation | # Samples<br>Illumina | # Samples<br>Nanopore | Predominant<br>Lineage |
| --- | --- | --- | --- |
| T12058C | 20 | 7 | B.1.177 |
| A28254C | 16 | 0 | B.1.351 (Beta) |
| C10789T | 14 | 2 | B.1.375 |
| G29692T | 10 | 2 | B.1.1.284 |
| C25904T | 10 | 1 | B.1.351 (Beta) |
| A24389C | 0 | 32 | B.1.1.7 (Alpha) |
| C23271A | 4 | 23 | B.1.1.7 (Alpha) |
| G24914C | 4 | 22 | B.1.1.7 (Alpha) |
| T8022G | 0 | 18 | B.1.617.2 (Delta) |
| C23709T | 3 | 16 | B.1.1.7 (Alpha) |

Two mutations preferentially detected by Illumina sequencing, A28254C and C25904T, were predominantly found in samples containing B.1.351 (Beta) lineage. The Beta variant was present in 50% of samples containing mutation A28254C at the time of sampling, but has since decreased to 20% of samples as the mutation is present in currently circulating variants. Mutation C25904T is considered a signature mutation of the Beta variant, though it was present in a number of other variants circulating at the time of sampling. The Beta variant was present in 64% of samples containing this variant at the time of sampling, and has since decreased in presence to 41%, while the mutation itself is still present in variants today, though largely decreased. Of the samples containing mutation T12058C, 70% contained the B.1.177 lineage at the time of sequencing, a variant that peaked in transmission in late 2020. Currently, multiple lineages contain this mutation, including some circulating today, with no significantly predominant lineage. The B.1.375 lineage was most prevalent in samples containing mutation C10789T, with a presence in 33% of samples at the time of sampling. Though this variant reached peak transmissibility in late 2020, the mutation is still present in a number of variants today. Samples containing mutation G29692T had a predominant lineage of B.1.1.284 at 38%, though the lineage B.1.1.176 was also highly present at 37%, at the time of sampling. Overall, however, the B.1.1.284 lineage has increased its predominance with a presence in 44% of all samples, while B.1.1.176 is only present in 10% of samples. This mutation is still present in circulating variants, but at a decreased presence compared to time of sampling.

Of the 5 mutations analysed that were preferentially detected by nanopore sequencing, three are considered signature mutations of the B.1.1.7 (Alpha) variant: C23271A, G24914C, and C23709T. For all samples containing any of these mutations, the Alpha variant was the predominant lineage, both at time of sampling and overall, with a 97-98% presence. Following this, the presence of these mutations is similar to the presence of the Alpha variant in the population and thus is non-existent today. The Alpha variant was also predominant in samples containing mutation A24389C, with a 50% presence at the time of sampling, which has decreased slightly to 39% to date. Like the three signature mutations, the presence of this mutation in samples decreased proportionately with the presence of the Alpha variant. The mutation T8022G has been sequenced in a low number of samples overall, with the majority of them containing the B.1.617.2 (Delta) lineage. At the time of sampling, the Delta lineage was present in 22% of samples containing the mutation, and has since risen to 34% presence.

### 3.5. Identifying Co-occurrence of Mutations within the Same Amplicon

To compare the sequencing rate of co-occurrence of mutations between both sequencing methods, we focused on signature mutations of the Alpha variant as this variant contains a number of co-occurrences in multiple amplicons (Table S3). We presented the number of co-occurrences compared to the overall frequency of the mutations within each dataset to determine the occurrence rate of these mutations (Figure 4). There were 5 amplicons in which co-occurrence of mutations occurs in the Alpha variant: amplicons 77, 78, 92, 93, and 95. In all amplicons, Nanopore detected these co-occurrences at an equal or higher frequency than in the equivalent samples sequenced by Illumina. This difference was significant in 3 amplicons: amplicon 77 (p = 0.0018), amplicon 78 (p = 0.0076), amplicon 93 (p = 0.049). However, both Illumina and Nanopore detected the co-occurrence of mutations in almost all samples in which the lesser frequent mutation in the amplicon was present.

**Figure 4.**
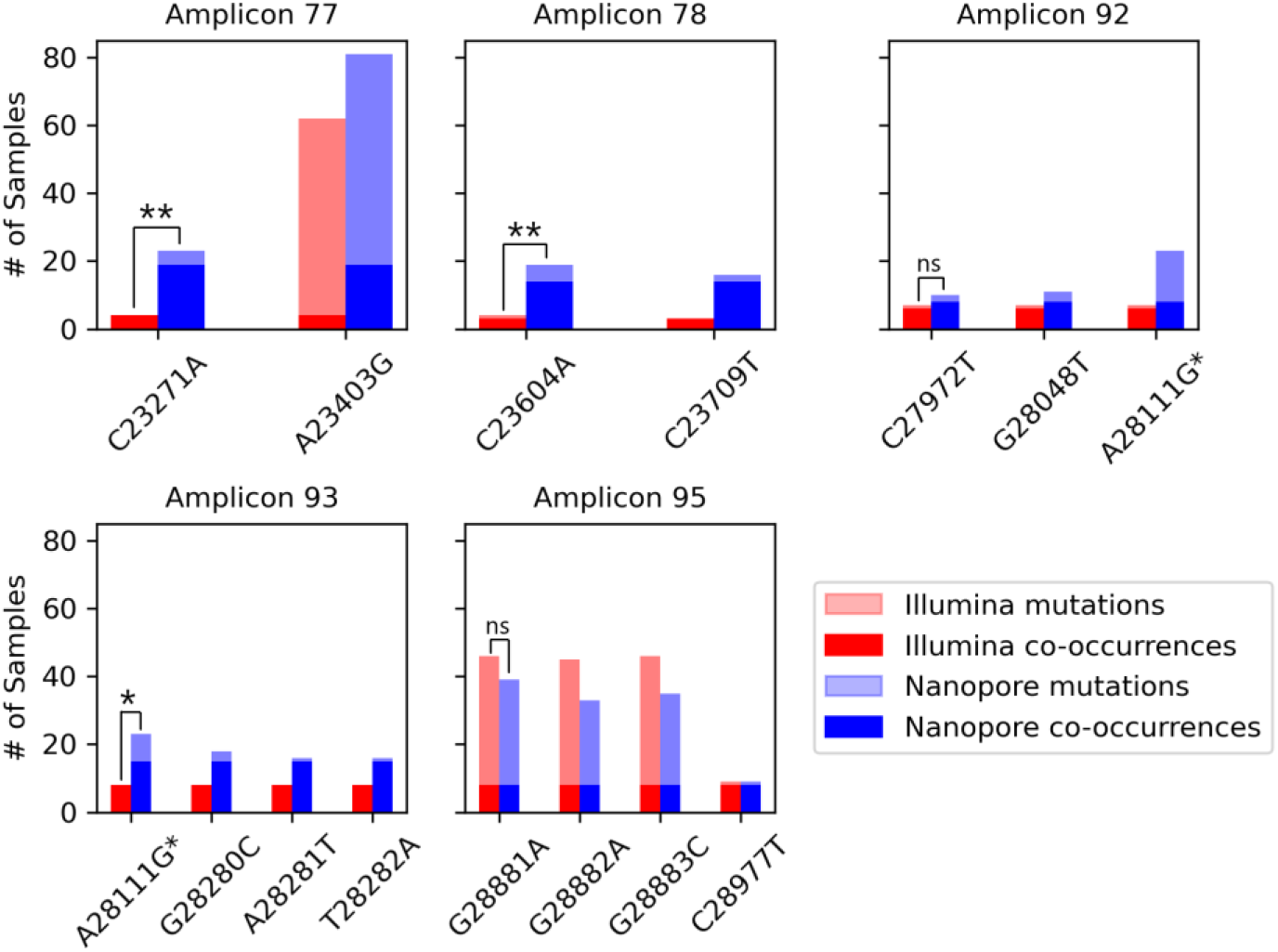
Co-occurrence of B.1.1.7 (Alpha) signature mutations within the same amplicon. Dark red and blue bars indicate the number of samples containing the co-occurrence in the Illumina and Nanopore datasets, respectively, while light bars show the overall frequency of the mutation. *Mutation A28111G is located in both Amplicon 92 and 93. Chi-squared test: **: p < 0.01, **: p < 0.05, ns: p > 0.05.

Looking at mutations occurring sequentially, GAT28280CTA and GGG28881AAC, all three single nucleotide changes occur at equal or similar frequency compared to the non-sequential signature mutation in the amplicon. Mutations at positions 28280-28282 occurred in 8 samples sequenced by Illumina, in which a co-occurrence without mutation A28111G was detected in all 8. Of the 7 samples including the non-sequential mutation, the detection rate remained at 100%. Conversely, mutation A28111G was detected in a higher number of Nanopore samples than mutations in positions 28280-28282 (23 samples vs 16 samples (28281 and 28282) and 18 samples (28280)), of which only 15 samples were detected with the co-occurrence both with and without A28111G. In amplicon 95, mutations C28977T was sequenced at a much lower frequency than mutations at positions 28881-28883 in both datasets. Mutations at position 28881-28883 were the only Alpha signature mutations sequenced at a higher frequency in the Illumina sample set than the Nanopore sample set. Both sequencing methods detect the co-occurrence of the three mutations in all samples in which the lowest frequent mutation was present, though all three mutations occurred in a very similar frequency.

## 4. Discussion

Advances in next generation sequencing have provided a breakthrough in detecting SARS-CoV-2 and tracking evolving variant lineages during the COVID-19 pandemic [8]. Using various methods, we have compared the efficacy of Illumina and Nanopore sequencing at detecting variants and the advantages of using one sequencing method over the other. Major differences in the reads between the Nanopore and Illumina datasets include a higher depth and accuracy using Illumina sequencing, which is to be expected for typical RNA sequencing [19]. This meant a greater amount of filtering was needed on samples sequenced by Nanopore in order to make a more meaningful comparison. However, Illumina’s reads did not span the length of the amplicons used for sequencing, which means that inference is needed to link single mutations in order to detect variants. Nanopore sequencing provides more direct evidence of variants in which its mutations occur along the same amplicon, as is the case for B.1.1.7. Looking at variants present in the wastewater samples, as well as key mutations differing between the two sequencing methods, we were able to determine a pattern in the differences between Illumina and Nanopore sequencing. Using this, we present a workflow that integrates data from the two sequencing technologies to get more comprehensive detection of SARS-CoV-2 variants in wastewater.

Using signature and marker mutations, we were able to identify VOCs and VOIs present within the Illumina and Nanopore dataset. A robust tool used during the pandemic is Freyja, which recovers relative lineage abundances from mixed SARS-CoV-2 samples such as wastewater [37]. For example, Freyja was used to perform VOC per sample analysis on wastewater samples as part of an environmental surveillance study in Malawi, which has limited COVID-19 testing capacity and no formal sewage systems [42]. Due to the lower coverage spread in our wastewater samples, Freyja identified a high number of “other” variants, which made it difficult to clearly visualise variant abundances. However, we were able to utilise the tool to confirm the presence of VOCs that we had identified using signature mutations. Our method also allowed us to focus on the co-occurrence of mutations within amplicons, as well as mutations with a high discordance in frequency between datasets. Nanopore sequencing detected 5 VOCs/VOIs at an equal or higher frequency than in Illumina samples: Alpha, Gamma, Zeta, and Eta, and Delta. Illumina was able to detect these variants with some overlap in samples, thereby increasing the frequency of samples containing a variant when taking into account either sequencing method. Conversely, Illumina detected 4 VOCs/VOIs at a higher frequency than in Nanopore sequenced samples (Beta, Theta, Lambda and Mu), as Nanopore sequencing did not detect these variants in any samples. Alpha, Gamma and Delta variants were the most prevalent in the area, while the Beta variant detected by Illumina had a low presence compared to the variants detected by Nanopore sequencing [43]. Aside from the Alpha and Beta variant, the low frequency of samples containing variants led to an insignificant difference between datasets. Detection of low frequent variants can be due to the high accuracy and higher coverage of Illumina sequencing. We have shown that Illumina detects individual mutations of lesser frequent and relatively unknown variants at a higher frequency than Nanopore sequencing due to higher accuracy and sequencing depth, while Nanopore preferentially detects mutations that are most predominant in highly frequent variants such as the Alpha variant.

The co-occurrence of mutations in the same amplicon is a more robust indicator of the presence of a variant in wastewater samples [39]. As Nanopore is able to sequence reads spanning the entire amplicon, the frequency of mutations can be used to provide more direct evidence of variants. Our data shows a similar frequency in mutations in the same amplicon in many instances, and co-occurrence with other signature mutations occurred in a high number of samples where a mutation was present. While the use of co-occurrences is not useful for all lineages, particularly in those where mutations are not close enough to be sequenced in the same amplicon, it can provide earlier detection for variants where this occurs. Similarly, a high frequency of co-occurrences within a sample can provide a benchmark for new lineages. The increased frequency of co-occurrences in Nanopore compared to Illumina highlights its advantages and can explain why it detects Alpha better. While the presence of mutations in the same amplicon across the population may be inferred to have the presence of a variant, it is only through long read sequencing that we can resolve multiple mutations in the same read.

We present a workflow that combines Illumina and Nanopore wastewater sequencing data, improving the detection of SARS-CoV-2 variants (Figure 5). As shown above, both sequencing technologies detected variants at different frequencies depending on their abundance in the wastewater as well as specific mutations in the variant. By integrating both datasets, we can use the strengths of each sequencing method to increase the number of variants found within a sample. A majority of the workflow remains the same as workflows involving SARS-CoV-2 variant analysis using one sequencing technology. The same wastewater collection protocol can be used, as well as RNA extraction and SARS-CoV-2 amplification. Separate amplicon PCR products from the same wastewater sample will go through library preparation with either Illumina and Nanopore protocols, and will therefore be sequenced with the respective instruments. Downstream processing of raw sequencing data, such as trimming and alignment, and mutational calling will occur separately between Illumina and Nanopore datasets, using readily available tools. Mutations from each dataset can then be integrated into one dataset, and may be filtered as a group depending on quality control samples and background noise, which was particularly present in Nanopore samples. Finally, the integrated dataset can be used for SARS-CoV-2 variant analysis using the usual tools developed during the pandemic.

**Figure 5.**
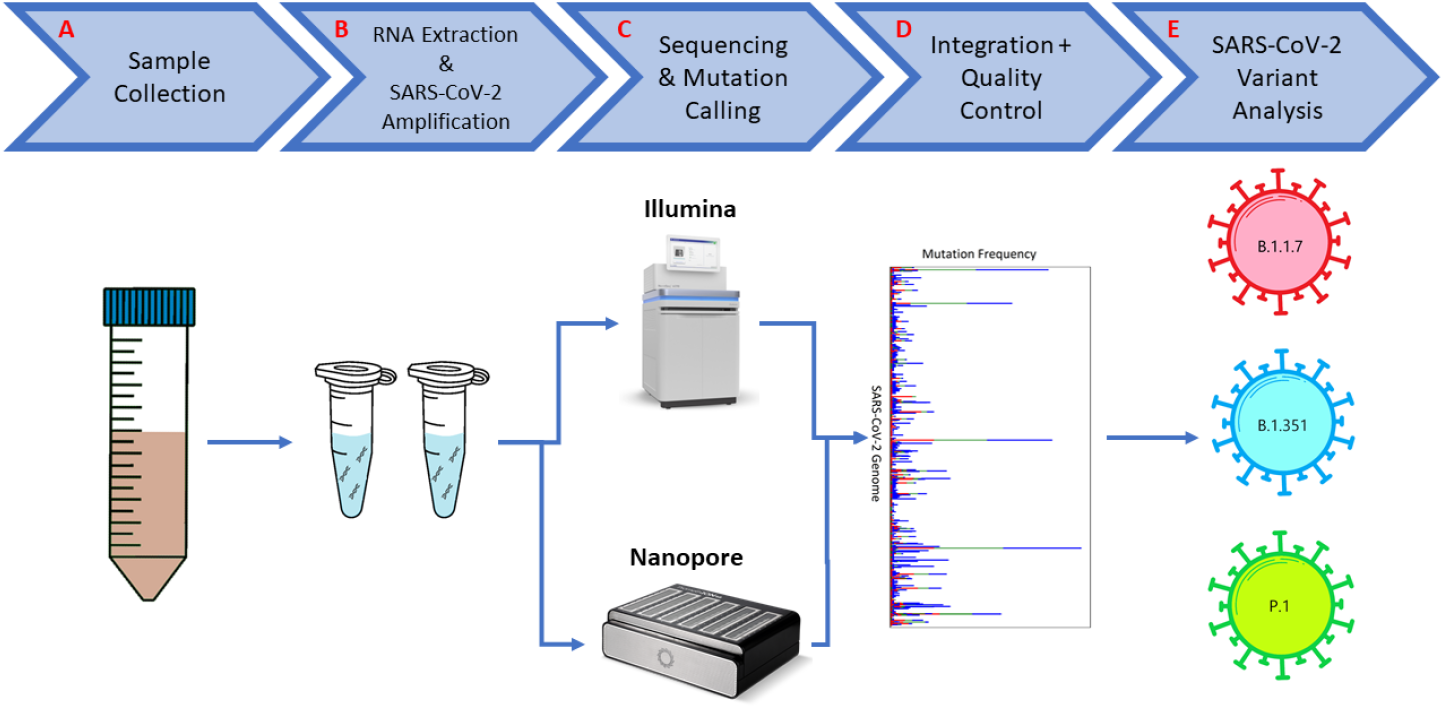
General workflow to integrate Illumina and Nanopore wastewater sequencing data. Steps include: A) collection of wastewater from target location; B) RNA extraction from wastewater and amplification of the SARS-CoV-2 genome; C) sequencing of the prepared sample on both Illumina and Nanopore instruments, followed by downstream processing and mutation calling using their respective tools; D) combining detected mutations from both datasets, with background noise removal; E) SARS-CoV-2 variant analysis using combined dataset.

When tracking the spread and evolution of SARS-CoV-2 variants using wastewater surveillance, Nanopore lacked the accuracy and depth to provide a full picture when compared to Illumina sequencing. While the sequencing turnaround is quicker, the lack of detection of certain variants can lead to a lag in public health response. Improved single read accuracy in Nanopore sequencing, as indicated using the recently released R10.4 chemistry, may improve detection of less frequent variants and detection of unknown variants. The current Nanopore chemistry allows for over 99% modal read accuracy, corresponding to a Phred score of Q21, while the chemistry used in this study has a lower Phred score of Q17 [44]. Although Nanopore sequencing is still limited by its lower throughput compared to Illumina, Oxford Nanopore is continuously improving their high throughput devices such as the PromethION. This study was also limited by the availability of data. Analysis was completed on basecalled Nanopore data as the raw signal data was unavailable. As there have been vast improvements in basecalling technology from Nanopore, using newer basecalling technology on the raw data may also lead to improved single read accuracy with increased performance [45,46]. Furthermore, looking at the raw signal allows us to analyse samples without the bias of a basecaller, using tools such as nanopolish [47,48].

While analysis was completed on samples taken at the height of the pandemic, during a peak in a major VOC, future studies may be done during periods of low virulence. This may be able to further differentiate the sequencing methods in detecting new mutations and variants without a known lineage. Our results show that combining short and long read sequencing improves detection of variant lineages in mixed population samples by providing earlier evidence of variants and increased detection of unknown lineages.

## Supporting information

Supplementary Analysis Scripts

## Data Availability

Raw sequencing data presented in the study are openly available in the NCBI database under BioProject ID PRJNA1127571 (https://www.ncbi.nlm.nih.gov/bioproject/1127571).

## Author Contributions

Conceptualization, S.K.L., P.V., S.D., R.D., B.J.S., D.F., J.R. and T.P.S.; methodology, G.J. and T.P.S.; software, G.J. and A.N.; validation, G.J., J.L. and J.H.G.; formal analysis, G.J.; investigation, G.J., J.L., S.J.R., S.C.A., S.L., S.C., A.T., F.S.Q., E.G. and M.H.; resources, G.J., J.L., J.H.G., S.J.R., S.C.A., A.N., A.T., F.S.Q., T.M., E.G., M.H., S.K.L., P.V., S.D., R.D., B.J.S., D.F., J.R. and T.P.S.; data curation, G.J., J.H.G., S.C.A., A.T., F.S.Q., T.M., E.G., M.H. and E.M.; writing—original draft preparation, G.J.; writing—review and editing, G.J., J.L., S.C.A., P.V., S.D., R.D., B.J.S., D.F., J.R. and T.P.S.; visualization, G.J.; supervision, J.R. and T.P.S.; project administration, J.R. and T.P.S.; funding acquisition, S.K.L., P.V., S.D., R.D., B.J.S., D.F., J.R. and T.P.S. All authors have read and agreed to the published version of the manuscript.

## Institutional Review Board Statement

Not applicable.

## Informed Consent Statement

Not applicable.

## Acknowledgments

We acknowledge the bioinformatics support from the Canadian Center for Computational Genomics (C3G), a platform at the McGill Genome Centre and Victor Phillip Dahdaleh Institute of Genomic Medicine. This research was supported in part through the computational resources and services provided by Advanced Research Computing at the University of British Columbia.

## References

1. Amicone M Borges, Alves MJ, Isidro J, Ze-Ze L, Duarte S, Vieira L, et al. Mutation rate of SARS-CoV-2 and emergence of mutators during experimental evolution. Evol Med Public Health 2022, 10(1), 142–155. doi: 10.1093/emph/eoac010

2. Updated working definitions and primary actions for SARS-CoV-2 variants. Available online: https://www.who.int/publications/m/item/updated-working-definitions-and-primary-actions-for--sars-cov-2-variants (accessed 30 January 2024)

3. Rambaut A, Holmes EC, O’Toole Á, Hill V, McCrone JT, Ruis C, du Plessis L & Pybus OG. A dynamic nomenclature proposal for SARS-CoV-2 lineages to assist genomic epidemiology. Nat Microbiol. 2020, 5(11), 1403–1407. doi: 10.1038/s41564-020-0770-5

4. Polo D, Baluja-Quintela M, Corbishley A, Jones DL, Singer AC, Graham DW & Romalde JL. Making waves: Wastewater-based epidemiology for COVID-19 – approaches and challenges for surveillance and prediction. Water Res. 2020, 186, 116404. doi: 10.1016/j.watres.2020.116404

5. Xiao A, Wu F, Bushman M, Zhang J, Imakaev M, Chai PR, Duvallet C, et al. Metrics to relate COVID-19 wastewater data to clinical testing dynamics. medRxiv 2021. doi: 10.1101/2021.06.10.21258580

6. Kallem P, Hegab HM, Alsafar H, Hasan SW & Banat F. SARS-CoV-2 detection and inactivation in water and wastewater: review on analytical methods, limitations and future research recommendations. Emerg Microbes Infect. 2023, 12(2), 2222850. doi: 10.1080/22221751.2023.2222850

7. Chen X, Kang Y, Luo J, Pang K, Xu X, Wu J, Li X & Jin S. Next-Generation Sequencing Reveals the Progression of COVID-19. Front. Cell. Infect. Microbiol. 2021, 11, 632490. doi: 10.3389/fcimb.2021.632490

8. John G, Sahajpal NS, Mondal AK, Ananth S, Williams C, Chaubey A, Rojiani AM & Kolhe R. Next-Generation Sequencing (NGS) in COVID-19: A Tool for SARS-CoV-2 Diagnosis, Monitoring New Strains and Phylodynamic Modeling in Molecular Epidemiology. Curr Issues Mol Biol 2021, 43(2), 845–867. doi: 10.3390/cimb43020061

9. Chiara M, D’Erchia AM, Gissi C, Manzari C, Parisi A, Resta N, Zambelli F, et al. Next generation sequencing of SARS-CoV-2 genomes: challenges, applications and opportunities. Brief Bioinform. 2020, bbaa297. doi: 10.1093/bib/bbaa297

10. Slatko BE, Gardner AF & Ausubel FM. Overview of Next Generation Sequencing Technologies. Curr Protoc Mol Biol 2018, 122(1), e59. doi: 10.1002/cpmb.59

11. Explore Illumina sequencing technology. Available online: https://www.illumina.com/science/technology/next-generation-sequencing/sequencing-technology.html (accessed 30 January 2024)

12. Nanopore DNA sequencing. Available online: https://nanoporetech.com/applications/dna-nanopore-sequencing (accessed 30 January 2024)

13. Szoboszlay M, Schramm L, Pinzauti D, Scerri J, Sandionigi A & Biazzo M. Nanopore is Preferable over Illumina for 16S Amplicon Sequencing of the Gut Microbiota When Species-Level Taxonomic Classification, Accurate Estimation of Richness or Focus on Rare Taxa Is Required. Microorganisms 2023, 11(3), 804. doi: 10.3390/microorganisms11030804

14. Linde J, Brangsch H, Holzer M, Thomas C, Elschner MC, Melzer F & Tomasco H. Comparison of Illumina and Oxford Nanopore Technology for genome analysis of Francisella tularensis, Bacillus anthracis, and Brucella suis. BMC Genomics 2023, 24(1), 258. doi: 10.1186/s12864-023-09343-z

15. Pecman A, Adams I, Gutierrez-Aguirre I, Fox A, Boonham N, Ravnikar M & Kutnjak D. Systematic Comparison of Nanopore and Illumina Sequencing for the Detection of Plant Viruses and Viroids Using Total RNA Sequencing Approach. Front Microbiol. 2022, 13, 883921. doi: 10.3389/fmicb.2022.883921

16. Stoler N & Nekrutenko A. Sequencing error profiles of Illumina sequencing instruments. NAR Genomics and Bioinformatics 2021, 3(1), lqab019. doi: 10.1093/nargab/lqab019.

17. Wang Y, Zhao Y, Bollas A, Wang Y & Au KF. Nanopore sequencing technology, bioinformatics and applications. Nature Biotechnology 2021, 39, 1348–1365. doi: 10.1038/s41587-021-01108-x

18. Tshiabuila D, Giandhari J, Pillay S, Ramphal U, Ramphal Y, Maharaj A, Anyaneji UJ, et al. Comparison of SARS-CoV-2 sequencing using the ONT GridION and the Illumina MiSeq. BMC Genomics 2022, 23, 319. doi: 10.1186/s12864-022-08541-5

19. Carbo EC, Mourik K, Boers SA, Munnink BO, Nieuwenhuijse D, Jonges M, Welkers MRA, et al, A comparison of five Illumina, Ion Torrent and nanopore sequencing technology-based approaches for whole genome sequencing of SARS-CoV-2. European Journal of Clinical Microbiology and Infectious Diseases 2023, 42, 701–713. doi: 10.1007/s10096-023-04590-0

20. Garcia-Pedemonte D, Carcereny A, Gregori J, Quer J, Garcia-Cehic D, Guerrero L, Cereto-Massague A, et al. Comparison of Nanopore and Synthesis-Based Next-Generation Sequencing Platforms for SARS-CoV-2 Variant Monitoring in Wastewater. Int J Mol Sci 2023, 24(24), 17184. doi: 10.3390/ijms242417184

21. D’Aoust PM, Graber TE, Mercier E, Montpetit D, Alexandrov I, Neault N, Baig AT, et al. Catching a resurgence: Increase in SARS-CoV-2 viral RNA identified in wastewater 48h before COVID-19 clinical tests and 96h before hospitalizations. Science of the Total Environment 2021, 770, 145319. doi: 10.1016/j.scitotenv.2021.145319

22. Hovi T, Stenvik M, Partanen H & Kangas A. Poliovirus surveillance by examining sewage specimens. Quantitative recovery of virus after introduction into sewerage at remote upstream location. Epidemiology & Infection 2001, 127(1), 101–106. doi: 10.1017/S0950268801005787

23. Petterson S, Grondahl-Rosado R, Nilsen V, Myrmel M & Robertson LJ. Variability in the recovery of a virus concentration procedure in water: Implications for QMRA. Water Research 2015, 87, 79–86. doi: 10.1016/j.watres.2015.09.006

24. Schang C, Crosbie ND, Nolan M, Poon R, Wang M, Jex A, John N, et al. Passive Sampling of SARS-CoV-2 for Wastewater Surveillance. Inviron. Sci. Technol 2021, 55(15), 10432–10441. doi: 10.1021/acs.est.1c01530

25. ARTIC Network – SARS-CoV-2. Available online: https://artic.network/ncov-2019 (accessed on 20 November 2023)

26. Martin M. Cutadapt Removes Adapter Sequences From High-Throughput Sequencing Reads. EMBnet.journal 2011, 17(1), 10–12. doi: 10.14806/ej.17.1.200

27. Li H. Aligning sequence reads, clone sequences and assembly contigs with BWA-MEM. arXiv 2013. doi: 10.48550/arXiv.1303.3997

28. Tarasov A, Vilella AJ, Cuppen E, Nijman IJ & Prins P. Sambamba: fast processing of NGS alignment formats. Bioinformatics 2015, 31(12), 2032–2034. doi: 10.1093/bioinformatics/btv098

29. Grubaugh ND, Gangavarapu K, Quick J, Matteson NL, Goes De Jesus J, Main BJ, Tan AL, et al. An amplicon-based sequencing framework for accurately measuring intrahost virus diversity using PrimalSeq and iVar. Genome Biology 2019, 20, 8. doi: 10.1186/s13059-018-1618-7

30. Li, H. A statistical framework for SNP calling, mutation discovery, association mapping and population genetical parameter estimation from sequencing data. Bioinformatics 2011, 27(21), 2987–2993. doi:10.1093/bioinformatics/btr509

31. Garrison E & Marth G. Haplotype-based variant detection from short-read sequencing. arXiv 2012. doi: 10.48550/arXiv.1207.3907

32. Nanoporetech/medaka. Available online: https://github.com/nanoporetech/medaka (accessed on 24 November 2023)

33. Accukit SARS-CoV-2. Available online: https://accugenomics.com/accukit-sars-cov-2/ (accessed on 24 November 2023)

34. Li, H. Minimap2: pairwise alignment for nucleotide sequences. Bioinformatics 2018, 34, 3094–3100. doi:10.1093/bioinformatics/bty191

35. CoV-lineages/constellations. Available online: https://github.com/cov-lineages/constellations (accessed on 24 November 2023)

36. N’Guessan A, Tsitouras A, Sanchez-Quete F, Goitom E, Reiling SJ, Galvez JH, Nguyen TL, et al. Detection of prevalent SARS-CoV-2 variant lineages in wastewater and clinical sequences from cities in Quebec, Canada. medRxiv 2022. doi:10.1101/2022.02.01.22270170

37. Karthikeyan S, Levy JI, De Hoff P, Humphry G, Birmingham A, Jepsen K, Farmer S, et al. Wastewater sequencing reveals early cryptic SARS-CoV-2 variant transmission. Nature 2022, 609, 101–108. doi: 10.1038/s41586-022-05049-6

38. Chen C, Nadeau S, Yared M, Voinov P, Ning X, Roemer C & Stadler T. CoV-Spectrum: Analysis of globally shared SARS-CoV-2 data to Identify and Characterize New Variants. Bioinformatics 2021, 38(6), 1735–1737 doi:10.1093/bioinformatics/btab856

39. Jahn K, Dreifuss D, Topolsky I, Kull A, Ganesanandamoorthy P, Fernandez-Cassi X, Banziger C, et al. Early detection and surveillance of SARS-CoV-2 genomic variants in wastewater using COJAC. Nature Microbiology 2022, 7, 1151–1160. doi:10.1038/s41564-022-01185-x

40. Van Rossum G & Drake FL. Python 3 Reference Manual; CreateSpace: Scotts Valley CA, 2009

41. Quality Scores for Next-Generation Sequencing. Available online: https://www.illumina.com/documents/products/technotes/technote_Q-Scores.pdf (accessed on 4 December 2023)

42. Barnes KG, Levy JI, Gauld J, Rigby J, Kanjerwa O, Uzzell CB, Chilupsya C, et al. Utilizing river and wastewater as a SARS-CoV-2 surveillance tool in settings with limited formal sewage systems. Nat Commun 2023, 14, 7883. doi: 10.1038/s41467-023-43047-y

43. Tracking variants of the novel coronavirus in Canada. Available online: https://www.ctvnews.ca/health/coronavirus/tracking-variants-of-the-novel-coronavirus-in-canada-1.5296141 (accessed 23 January 2024)

44. Ni Y, Liu X, Simeneh ZM, Yang M & Li R. Benchmarking of Nanopore R10.4 and R9.4.1 flow cells in single-cell whole-genome amplification and whole-genome shotgun sequencing. Computational and Structural Biotechnology Journal 2023, 21, 2352–2364. doi: 10.1016/j.csjb.2023.03.038

45. Nanoporetech/dorado. Available online: https://github.com/nanoporetech/dorado (accessed 23 January 2024)

46. Benchmarking the Oxford Nanopore Technologies basecallers on AWS. Available online: https://aws.amazon.com/blogs/hpc/benchmarking-the-oxford-nanopore-technologies-basecallers-on-aws/ (accessed 23 January 2024)

47. Ferguson S, McLay T, Andrew RL, Bruhl JJ, Schwessinger B, Borevitz J & Jones A. Species-specific basecallers improve actual accuracy of nanopore sequencing plants. Plant Methods 2022, 18, 137. doi: 10.1186/s13007-022-00971-2

48. jts/nanopolish. Available online: https://github.com/jts/nanopolish (accessed 23 January 2024)

