## Supplementary Analysis Scripts for "Combining short and long read sequencing technologies to identify SARS-CoV-2 variants in wastewater": SARS_WW_CoocAmp.py

```
"""
SARS-CoV-2 Wastewater Data Comparison - Nanopore vs Illumina

List co-occurrence of mutations within same amplicon
"""
import os 
import itertools

def main():
    with open('C:/Scripts/SARS_WW/SARS_WW-mut_cooccurrence.txt','w') as myfile:
        myfile.write('co-occurrence\tmatching\tillumina_only\tnanopore_only\n') 
    
    ## List of amplicons
    amplicon_list = []
    with open('C:/Scripts/SARS_WW/nCoV-2019_V3.bed') as myfile:
        all_data = myfile.read().splitlines()
        for x in range(1,99):
            for line in all_data:
                data = line.split()
                primer_num = data[3].split('_')[1]
                if int(primer_num) == x:
                    if data[5] == '+':
                        start = data[1]
                    if data[5] == '-':
                        end = data[2]
            amplicon_list.append([start,end])
    
    ## Get list of all samples that can be compared
    sample_list = []
    with open('C:/Scripts/SARS_WW/sample.txt') as myfile:
        for line in myfile:          
            sample_list.append(line.rstrip())
            
    ## Determine control snps to remove
    noisy_muts = []
    for x in sample_list:
        if 'Ctrl' in x:   
        ## Get Illumina variant file based on sample name
            illumina_file = get_ifile(x)
        ## Get Illumina variants
            illumina_var = get_idata(illumina_file)      
        ## Get Nanopore medaka file based on flowcell, barcode
            nanopore_file = get_npfile(x)
        ## Get Nanopore variants
            nanopore_var = get_npdata(nanopore_file)
        ## Compare variants
            common_muts, illumina_remove, nanopore_remove = match_muts(illumina_var, nanopore_var)
        ## create list of 'noisy' SNPs (illumina and nanopore only)
            noisy_muts.extend(illumina_remove)
            noisy_muts.extend(nanopore_remove)

    ## Add nanopore muts under threshold to noisy muts
    with open('C:/Scripts/SARS_WW/SARS_WW-mut_count_unfiltered.txt','r') as myfile:
        next(myfile)
        for line in myfile:
            mut, both, illumina, nanopore = line.split()[0], line.split()[1], line.split()[2], line.split()[3]
            if int(both) == 0 and int(illumina) == 0 and int(nanopore) < 10:
                noisy_muts.append(mut)
                
    co_muts = dict()            
    for x in sample_list:
    ## Get Illumina variant file based on sample name
        illumina_file = get_ifile(x)
    ## Get Illumina variants
        illumina_var = get_idata(illumina_file) 
        illumina_muts = [m for m in illumina_var if m not in noisy_muts]  
    ## Get Illumina co-muts
        illumina_comuts = find_comuts(illumina_muts, amplicon_list)
    ## Get Nanopore medaka files based on sample name
        nanopore_file = get_npfile(x)
    ## Get Nanopore variants
        nanopore_var = get_npdata(nanopore_file)
        nanopore_muts = [m for m in nanopore_var if m not in noisy_muts]  
    ## Get Nanopore co-muts
        nanopore_comuts = find_comuts(nanopore_muts, amplicon_list)
    ## Compare co-muts
        match_comuts = list(set(illumina_comuts) & set(nanopore_comuts))
        ill_only_comuts = list(set(illumina_comuts) - set(nanopore_comuts))
        np_only_comuts = list(set(nanopore_comuts) - set(illumina_comuts)) 
    ## Add to dictionary   
        for x in match_comuts:
            if x not in co_muts:
                co_muts[x] = [0,0,0]  
            co_muts[x][0] += 1
        for x in ill_only_comuts:
            if x not in co_muts:
                co_muts[x] = [0,0,0]
            co_muts[x][1] += 1
        for x in np_only_comuts:
            if x not in co_muts:
                co_muts[x] = [0,0,0]
            co_muts[x][2] += 1
    ## Write to file
    with open('C:/Scripts/SARS_WW/SARS_WW-mut_cooccurrence.txt','a') as myfile:
        for x in dict(sorted(co_muts.items(), key=lambda item: (item[1][0],item[1][1],item[1][2]), reverse=True)):
            myfile.write('{}\t{}\t{}\t{}\n'.format(x,co_muts[x][0],co_muts[x][1],co_muts[x][2]))

def get_ifile(sample):
    '''Get illumina TSV file from sample name.'''
    illumina_folder = "C:/Scripts/SARS_WW/illumina_variants"
    illumina_file = os.path.join(illumina_folder, sample+'.variants.tsv')
 
    return illumina_file

def get_idata(illumina_file):
    '''Get variants from Illumina TSV file.'''
    illumina_var = []
    with open(illumina_file) as myfile:
        next(myfile)
        for line in myfile:
            pos, ref, alt = line.split()[1], line.split()[2], line.split()[3]
            if '-' in alt:
                ref += alt[1:]
                alt = ref[0]
            if '+' in alt:
                alt = ref + alt[1:]
                
            var = '{}{}{}'.format(ref,pos,alt)
            illumina_var.append(var)
            
    return illumina_var      
    
def get_npfile(sample):
    '''Get medaka VCF file from sample name.'''
    nanopore_folder = "C:/Scripts/SARS_WW/nanopore_variants"
    nanopore_file = os.path.join(nanopore_folder, sample+'.vcf')

    return nanopore_file

def get_npdata(nanopore_file):
    '''Get variants from medaka VCF file.'''
    nanopore_var = []
    with open(nanopore_file) as myfile:
        for line in myfile:
            if line.startswith('#'): continue
            data = line.split()
            var = '{}{}{}'.format(data[3],data[1],data[4])
            ## split combined mutations
            if len(data[3]) > 1 and len(data[4]) > 1 and len(data[3]) == len(data[4]):
                for x in range(0, len(data[3])):
                    split_var = '{}{}{}'.format(data[3][x],str(int(data[1])+x),data[4][x])
                    nanopore_var.append(split_var)
            else:
                nanopore_var.append(var)
      
    return nanopore_var 

def match_muts(illumina_var, nanopore_var):
    '''From two variant lists: get common mutations, illumina only, nanopore only'''
    matches = list(set(illumina_var).intersection(nanopore_var))
    illumina_only = list(set(illumina_var).difference(nanopore_var))
    nanopore_only = list(set(nanopore_var).difference(illumina_var))
    
    return matches, illumina_only, nanopore_only 

def find_comuts(var_list, amplicon_list):
    '''From list of variants: find co-occurrence in same amplicon.'''
    co_muts = []
    all_pairs = list(itertools.combinations(var_list,2))
    for amp in amplicon_list:
        for pair in all_pairs:
            pos1 = ''.join(x for x in pair[0] if x.isdigit())
            pos2 = ''.join(x for x in pair[1] if x.isdigit())
            if int(amp[0]) <= int(pos1) <= int(amp[1]) and int(amp[0]) <= int(pos2) <= int(amp[1]) and int(pos1) != int(pos2):
                if int(pos1) > int(pos2):
                    co_muts.append((pair[1],pair[0]))
                else:
                    co_muts.append((pair[0],pair[1]))
    
    return co_muts 

def write_muts(comut_list):
    '''Format list of comuts -> [('mut1','mut2')]'''
    if not comut_list:
        return '-'
    else:
        comut_string = []
        for x in comut_list:
            comut_string.append('({},{})'.format(x[0],x[1]))
        mutations = ','.join(comut_string)
        return mutations    
    
if __name__ == "__main__":
    main()
```
