## Supplementary Analysis Scripts for "Combining short and long read sequencing technologies to identify SARS-CoV-2 variants in wastewater": SARS_WW_Lineage.py

```
"""
SARS-CoV-2 Wastewater Data Comparison - Nanopore vs Illumina

List relative abundance of VOC/VOIs among Illumina and Nanopore datasets
"""
import csv 
from collections import Counter
import re

def main():
    signature_dict = {}
    marker_dict = {}
    mutation_counter = Counter()
    
    ## Create dict of signature mutations
    with open('C:/Scripts/SARS_WW/signature_muts_VOC-VOI.csv','r') as myfile:
        data = csv.reader(myfile)
        next(data,None)
        for x in data:
            if x[0] not in signature_dict:
                signature_dict[x[0]] = []
            signature_dict[x[0]].append(x[1])
            mutation_counter.update([x[1]])

    ## Create dict of marker mutations
    for lineage in signature_dict:
        if lineage not in marker_dict:
            marker_dict[lineage] = []
        for mutation in signature_dict[lineage]:
            if mutation_counter[mutation] == 1:
                marker_dict[lineage].append(mutation)
   
    illumina_dict = {} 
    for lineage in signature_dict:
        illumina_dict[lineage] = []    
    nanopore_dict = {}
    for lineage in signature_dict:
        nanopore_dict[lineage] = []   
                              
    ## Compare samples to lineage mutations (3 signature, 1 marker minimum)                   
    with open('C:/Scripts/SARS_WW/SARS_WW-mut_comparison.txt', 'r') as myfile:
        next(myfile)
        for line in myfile:
            line = line.strip()
            if not line: continue
            data = line.split('\t')
            if 'Ctrl' in data[0]: continue
            
            match_muts = [] if '-' in data[1] else data[1].split(',')
            ill_muts = [] if '-' in data[2] else data[2].split(',')
            np_muts =  [] if '-' in data[3] else data[3].split(',')
            
            all_illumina = match_muts + ill_muts
            illumina_muts = []
            for x in all_illumina:
                illumina_muts.append("".join(re.split('(\d+)',x))[1:])
                
            all_nanopore = match_muts + np_muts
            nanopore_muts = []
            for x in all_nanopore:
                nanopore_muts.append("".join(re.split('(\d+)',x))[1:])

            for lineage in signature_dict:
                if len(set(signature_dict[lineage]) & set(illumina_muts)) > 2 and len(set(marker_dict[lineage]) & set(illumina_muts)) > 0:
                    illumina_dict[lineage].append(data[0])
                if len(set(signature_dict[lineage]) & set(nanopore_muts)) > 2 and len(set(marker_dict[lineage]) & set(nanopore_muts)) > 0:
                    nanopore_dict[lineage].append(data[0])  

    ## Count relative abundance of each lineage                
    lineage_counter = {}
    for lineage in signature_dict:
        lineage_counter[lineage] = [0,0,0,0]

        lineage_counter[lineage][0] = len(illumina_dict[lineage])
        lineage_counter[lineage][1] = len(nanopore_dict[lineage])
        lineage_counter[lineage][2] = len(set(illumina_dict[lineage]).intersection(nanopore_dict[lineage]))
        lineage_counter[lineage][3] = len(set(illumina_dict[lineage] + nanopore_dict[lineage]))
    
    ## Write to file
    with open('C:/Scripts/SARS_WW/SARS_WW-lineage_counts.txt', 'w') as myfile:
        myfile.write('lineage\tillumina\tnanopore\tboth\teither\n')
        for x in lineage_counter:
            myfile.write('{}\t{}\t{}\t{}\t{}\n'.format(x, lineage_counter[x][0],lineage_counter[x][1],lineage_counter[x][2],lineage_counter[x][3]))
   
         
if __name__ == "__main__":
    main()
```
