## Supplementary Analysis Scripts for "Combining short and long read sequencing technologies to identify SARS-CoV-2 variants in wastewater": SARS_WW_Mutations.py

```
"""
SARS-CoV-2 Wastewater Data Comparison - Nanopore vs Illumina

List and graph mutations present in Illumina and Nanopore datasets.
"""
import os 
import matplotlib.pyplot as plt
import matplotlib.patches as mpatches

def main():   
    with open('C:/Scripts/SARS_WW/SARS_WW-mut_count.txt','w') as myfile:
        myfile.write('mutation\tboth_datasets\tillumina_only\tnanopore_only\n') 
    
    ## Get list of all samples that can be compared
    sample_list = []
    with open('C:/Scripts/SARS_WW/sample.txt') as myfile:
        for line in myfile:          
            sample_list.append(line.rstrip())
    
    ## Determine control SNPs to remove
    noisy_muts = []
    for x in sample_list:
        if 'Ctrl' in x:     
        ## Get Illumina variant file based on sample name
            illumina_file = get_ifile(x)
        ## Get Illumina variants
            illumina_var = get_idata(illumina_file)   
        ## Get Nanopore medaka file based on sample name
            nanopore_file = get_npfile(x)    
        ## Get Nanopore variants
            nanopore_var = get_npdata(nanopore_file)
        ## Compare variants
            common_muts, illumina_remove, nanopore_remove = match_muts(illumina_var, nanopore_var)
        ## create list of 'noisy' SNPs (illumina and nanopore only)
            noisy_muts.extend(illumina_remove)
            noisy_muts.extend(nanopore_remove)
            
    mutations = dict()
    for x in sample_list:  
    ## Get Illumina variant file based on sample name
        illumina_file = get_ifile(x)
    ## Get Illumina variants
        illumina_var = get_idata(illumina_file)      
    ## Get Nanopore medaka files based on sample name
        nanopore_file = get_npfile(x)
    ## Get Nanopore variants
        nanopore_var = get_npdata(nanopore_file)
    ## Compare variants
        common_muts, illumina_only, nanopore_only = match_muts(illumina_var, nanopore_var)
    ## add each variant to dict
        for x in common_muts:
            if x not in mutations:
                mutations[x] = [0,0,0]
            mutations[x][0] += 1
        for x in illumina_only:
            if x not in mutations:
                mutations[x] = [0,0,0]
            mutations[x][1] += 1
        for x in nanopore_only:
            if x not in mutations:
                mutations[x] = [0,0,0]
            mutations[x][2] += 1
    
    ## remove noisy muts with nanopore_only < 10
    for x in mutations:
        if mutations[x][0] == 0 and mutations[x][1] == 0 and mutations[x][2] < 10:
            noisy_muts.append(x)
    noisy_muts = set(noisy_muts)
            
    for x in noisy_muts:
        del mutations[x]
        
    ## Write to file
    with open('C:/Scripts/SARS_WW/SARS_WW-mut_count.txt','a') as myfile:
        for x in dict(sorted(mutations.items(), key=lambda item: (item[1][0],item[1][1],item[1][2]), reverse=True)):
            myfile.write('{}\t{}\t{}\t{}\n'.format(x,mutations[x][0],mutations[x][1],mutations[x][2]))
            
    ## Graph mutation
    fig, (ax1,ax2,ax3) = plt.subplots(3,sharex=True,sharey=True)
    for y in mutations:
        pos = int(''.join(x for x in y if x.isdigit()))
        match = mutations[y][0]
        illumina = mutations[y][1]
        nanopore = mutations[y][2]
        ax1.bar(pos,illumina,color='r',width=100)
        ax1.bar(pos,match,color='forestgreen', bottom = illumina,width=100)
        ax1.bar(pos,nanopore,color='b', bottom = illumina+match,width=100)
        ax2.bar(pos,illumina+match,color='r',width=100)
        ax3.bar(pos,nanopore+match,color='b',width=100)
    
    ill_data = mpatches.Patch(color='r', label='Illumina')
    np_data = mpatches.Patch(color='b', label='Nanopore')
    both = mpatches.Patch(color='forestgreen',label='Both methods')
    ax1.legend(handles=[ill_data,np_data, both],loc='upper right',fontsize='x-small')    
    plt.savefig('C:/Scripts/SARS_WW/SARS_WW-mutations.png', dpi=300)


def get_ifile(sample):
    '''Get illumina TSV file from sample name.'''
    illumina_folder = "C:/Scripts/SARS_WW/illumina_variants"
    illumina_file = os.path.join(illumina_folder, sample+'.variants.tsv')
 
    return illumina_file

def get_idata(illumina_file):
    '''Get variants from Illumina TSV file.'''
    illumina_var = []
    with open(illumina_file) as myfile:
        next(myfile)
        for line in myfile:
            pos, ref, alt = line.split()[1], line.split()[2], line.split()[3]
            if '-' in alt:
                ref += alt[1:]
                alt = ref[0]
            if '+' in alt:
                alt = ref + alt[1:]
                
            var = '{}{}{}'.format(ref,pos,alt)
            illumina_var.append(var)
            
    return illumina_var      
    
def get_npfile(sample):
    '''Get medaka VCF file from sample name.'''
    nanopore_folder = "C:/Scripts/SARS_WW/nanopore_variants"
    nanopore_file = os.path.join(nanopore_folder, sample+'.vcf')

    return nanopore_file

def get_npdata(nanopore_file):
    '''Get variants from medaka VCF file.'''
    nanopore_var = []
    with open(nanopore_file) as myfile:
        for line in myfile:
            if line.startswith('#'): continue
            data = line.split()
            var = '{}{}{}'.format(data[3],data[1],data[4])
            ## split combined mutations
            if len(data[3]) > 1 and len(data[4]) > 1 and len(data[3]) == len(data[4]):
                for x in range(0, len(data[3])):
                    split_var = '{}{}{}'.format(data[3][x],str(int(data[1])+x),data[4][x])
                    nanopore_var.append(split_var)
            else:
                nanopore_var.append(var)
      
    return nanopore_var
  
def match_muts(illumina_var, nanopore_var):
    '''
    From two variant lists: get common mutations, illumina only, nanopore only
    '''
    matches = list(set(illumina_var).intersection(nanopore_var))
    illumina_only = list(set(illumina_var).difference(nanopore_var))
    nanopore_only = list(set(nanopore_var).difference(illumina_var))
    
    return matches, illumina_only, nanopore_only
    

if __name__ == "__main__":
    main()
```
